# Early-childhood dietary patterns associate with asthma and stool-plasma metabolomic signatures

**DOI:** 10.64898/2026.08.24.26361154

**Authors:** Yu Wei, Tinashe Chikowore, Scott T. Weiss, Yang-Yu Liu, Xu-Wen Wang

**Affiliations:** Channing Division of Network Medicine, Department of Medicine, Brigham and Women’s Hospital and Harvard Medical School, Boston, Massachusetts 02115, USA

## Abstract

**Background:** Childhood asthma has been linked to individual foods, nutrients, diet-quality scores, and broad dietary patterns, but specific early-childhood food co-consumption patterns and their microbial/metabolic correlates remain unclear.

**Objective:** To identify data-driven early-childhood dietary patterns associated with asthma/wheeze, evaluate prospective associations with age-6 asthma/wheeze, assess external support in NHANES, and characterize associated gut microbiome and stool/plasma metabolomic profiles.

**Methods:** We analyzed age-3 food frequency questionnaire data from children in the Vitamin D Antenatal Asthma Reduction Trial. Dietary patterns were derived from log-transformed, energy-residualized, standardized food-frequency variables using principal component analysis. Associations with age-3 asthma/wheeze were tested using covariate-adjusted logistic regression. Prospective associations were evaluated using age-6 asthma/wheeze as the outcome. Leading PC food-cluster proxies were evaluated in NHANES 2021-2023 among children aged 2-3 years, with sensitivity analyses in ages 2-5 and 2-8 years. Selected PCs were tested for associations with gut microbiome, stool metabolome, and plasma metabolome features.

**Results:** PC1 contrasted a sweet snack/fried-food pattern with a fruit/vegetable-rich pattern, whereas PC3 captured a processed meat/fried-food axis. PC3 showed the strongest positive association with age-3 asthma/wheeze (odds ratio per 1-SD increase, 1.42; P = 0.00109). Age-3 dietary PCs were prospectively associated with age-6 asthma/wheeze, with the overall PC set improving model fit in permutation testing (likelihood-ratio statistic = 20.3; empirical P = 0.033) among 394 cases and 397 controls. In NHANES children aged 2-3 years, the PC3 food-cluster proxy was positively associated with current asthma (odds ratio, 1.59; 95% confidence interval, 0.95-2.67). PC3 was also linked to gut microbial and stool/plasma metabolomic variation, including steroid sulfate, vitamin E-related, nucleoside-related, and lipid-related metabolites.

**Conclusions:** Early-childhood asthma/wheeze-associated dietary signals were better represented as food co-consumption patterns than isolated single-food effects. Age-3 dietary patterns were associated with concurrent and prospective asthma/wheeze, showed directionally consistent NHANES support, and were linked to microbiome and metabolomic variation.

## Introduction

Asthma remains one of the most common chronic diseases of childhood^1–3^ and continues to impose substantial morbidity on children, families, and health systems^4^. Recent U.S. surveillance data estimate that current asthma affects approximately 6% to 7% of children^5^, with persistent disparities by age, race, socioeconomic context^6^, and environmental exposure^7^. Although pharmacologic therapy can reduce symptoms and exacerbations^8,9^, prevention and early-life risk modification remain major clinical and public health goals^10–12^.

Diet is a plausible modifiable factor in asthma development and morbidity^13,14^. Prior epidemiologic studies have linked higher intake of fruits^15^, vegetables^16^, fish^17^, and Mediterranean-style dietary patterns with lower asthma or wheeze risk in some pediatric populations^18–20^, whereas Western-style^21^, fast-food^22^, or ultra-processed dietary patterns^23,24^ have often been hypothesized to increase risk^25,26^. However, evidence linking diet to pediatric asthma remains heterogeneous across populations, age windows, exposure definitions, and analytic strategies^27–29^. This inconsistency suggests that the relevant exposure may not be a single nutrient or food, but rather a higher-dimensional dietary structure embedded within social, behavioral, and biological contexts.

Previous diet–asthma studies have examined individual foods or nutrients as separate exposures. However, children do not consume foods in isolation. Dietary-pattern methods are therefore essential for studying diet in asthma, which summarize correlated food-intake variables into empirical axes of co-consumption that better reflect real-world eating behavior^30,31^. Principal component analysis has been widely used in nutritional epidemiology to derive dietary patterns from FFQ data^32^, providing a statistically efficient approach to summarize correlated food-intake variables into a smaller set of pattern scores that capture food co-consumption structure while reducing the multiple-testing burden and multicollinearity inherent in single-food analyses^30,33,34^. In asthma and respiratory epidemiology, PCA-based studies have identified broad patterns, or population-specific dietary patterns, and tested their associations with asthma, wheeze, lung function, asthma control, or related allergic phenotypes^35–37^. These studies suggested that diet–asthma associations may operate at the level of overall food-consumption patterns rather than isolated foods.

However, PCA-derived dietary patterns are interpreted primarily from food loadings and then analyzed as epidemiologic exposures^38,39^. The dietary principal component may represent a biologically meaningful dietary axis, but it may also capture cultural food combinations, socioeconomic patterning^40–42^, questionnaire structure, or reporting behavior^43,44^. Thus, food loadings and clinical associations alone are insufficient to determine whether an asthma-associated dietary pattern reflects an underlying biological pathway. It is critical to connect PCA-derived dietary patterns with molecular and microbial phenotypes that can biologically annotate these empirical food axes^45,46^. The gut microbiome and metabolome provide a plausible mechanistic bridge between diet and immune-mediated airway disease^47–49^. Early-life gut microbial dysbiosis has been associated with subsequent asthma risk^50–52^, and microbial metabolites, including short-chain fatty acids^52,53^, polyunsaturated fatty acid derivatives^54,55^, bile acids^56,57^, amino-acid derivatives^58,59^, and other small molecules^60^, may influence immune development and airway inflammation through the gut–lung axis^61^. Linking PCA-based dietary patterns to gut microbial composition, stool metabolomic profiles^62^, and circulating plasma metabolomic signatures^63^ can help distinguish dietary axes that are merely statistical summaries of co-consumption from those that may represent biologically relevant food–microbiome– metabolite programs involved in asthma development^64^.

Here, we identified year-3 dietary patterns in VDAART and tested the associations between dietary PCs and asthma/wheeze. We found a leading PC driven by confectionery foods. We externally evaluated NHANES food-cluster proxies for the leading asthma/wheeze-associated VDAART dietary PCs, including PC1, PC3, and PC9. Finally, we tested the three PCs separately for associations with gut microbiome, stool metabolome, and plasma metabolome features, providing biological annotation of the three dietary patterns.

## Methods

### Study design

We conducted an analysis of VDAART year-3 data to identify empirical dietary co-consumption patterns associated with asthma/wheeze and to characterize corresponding gut microbiome and metabolomic signatures (see **Figure 1**). We then performed external validation using NHANES 2021–2023 dietary recall and asthma questionnaire data. We derived dietary PCs from VDAART FFQ data, tested PC–asthma/wheeze association, externally evaluated food-cluster proxies in NHANES, and finally tested the VDAART PC1, PC3, and PC9 scores separately for associations with gut microbiome, stool metabolome, and plasma metabolome features.

**Figure 1.**
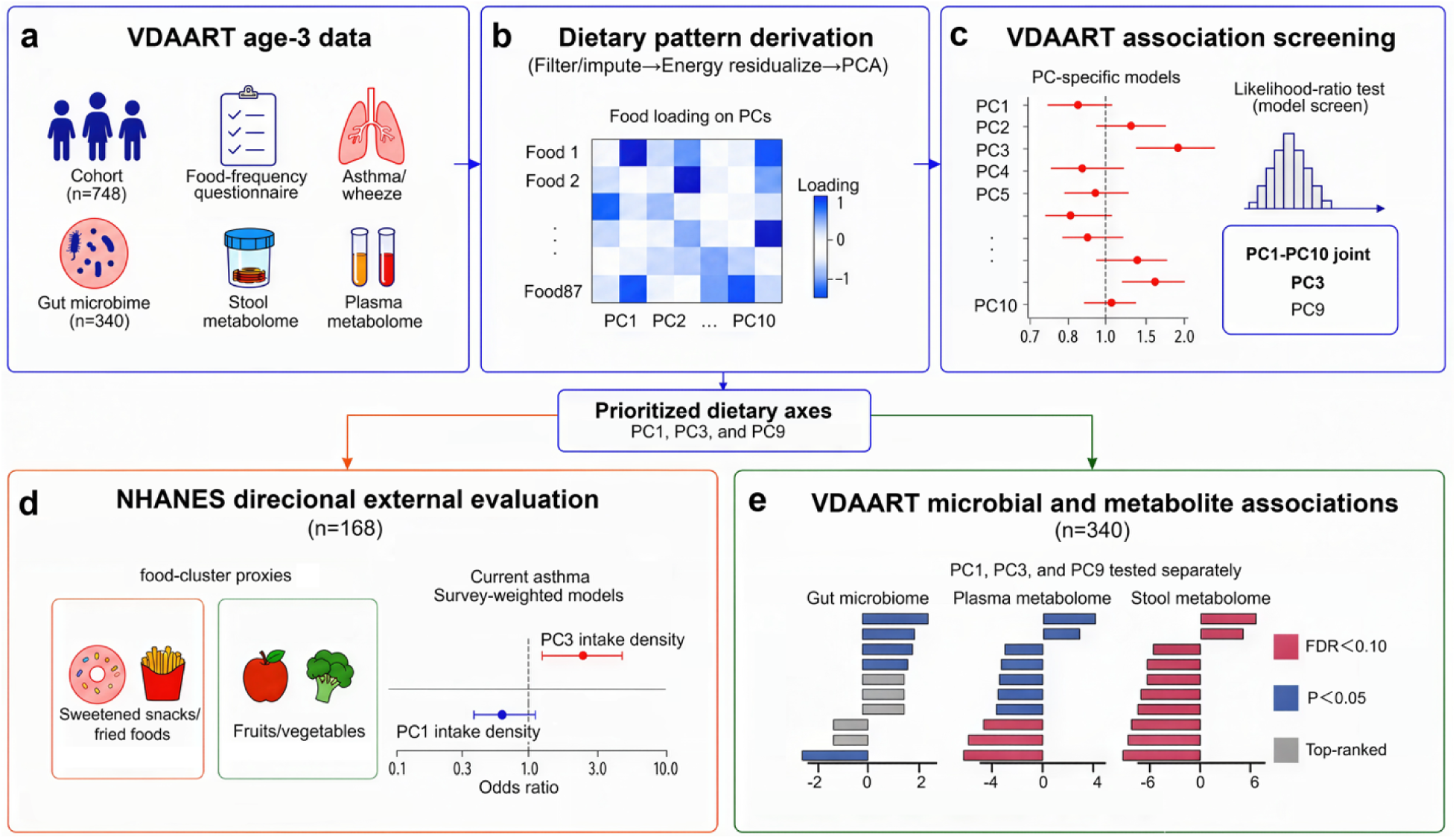
Study design and analytical workflow. **(a)** Age-3 VDAART data included dietary intake, asthma/wheeze outcomes, gut microbiome, stool metabolome, and plasma metabolome data. **(b)** Dietary data were processed and analyzed by principal component analysis to derive dietary patterns. **(c)** Dietary pattern scores were tested for associations with asthma/wheeze. **(d)** Food-cluster proxies of individual dietary axes that contributed most strongly to asthma were evaluated in NHANES. The main analysis included children aged 2–3 years, with ages 2–5 and 2–8 years used for sensitivity analyses. **(e)** Individual dietary axes showing the strongest association with asthma/wheeze were tested separately for associations with gut microbiome, stool metabolome, and plasma metabolome features.

### VDAART study population

VDAART is a randomized clinical trial designed to test whether prenatal vitamin D supplementation reduced asthma and allergy risk in offspring. Pregnant women aged 18–40 years with an estimated gestational age of 10–18 weeks were recruited from three clinical centers: Boston Medical Center, Washington University in St. Louis, and Kaiser Permanente Southern California. Details of the parent trial design and follow-up have been described previously^65^. In the present analysis, we included children with available year-3 FFQ data (87 food items in total), asthma/wheeze outcome data, and relevant covariates. Participants were excluded from specific analyses if they lacked the outcome, dietary PC score, total energy variable, or covariates required for that model. In the full VDAART offspring cohort with available asthma classification, 125 of 748 children had asthma. In the stool-metabolomics subset, 85 of 361 children had asthma^66^. The multi-omic analytic dataset used for diet– microbiome–metabolome integration included 340 children with age-3 FFQ, gut microbiome, stool metabolome, and plasma metabolome data.

### VDAART asthma/wheeze outcome

The primary VDAART outcome for this analysis was year-3 asthma/wheeze, defined from the available year-3 asthma/wheeze variable and coded as a binary outcome. Asthma assessment was based on maternal report of a physician diagnosis during the first three years of the child’s life. Because recent respiratory symptoms may better identify young children with clinically significant asthma, asthma/wheeze was defined using physician-diagnosed asthma together with recent wheeze or asthma-related symptoms when available^65^. Children were classified as cases if the year-3 asthma/wheeze variable indicated asthma or wheeze and as controls otherwise. The outcome was used only after dietary PCs were derived; asthma/wheeze status was not used to construct dietary patterns.

### FFQ processing and missingness handling

Year-3 child FFQ responses were converted to estimated intake frequency in times per day. Frequency categories were mapped to numeric daily frequencies, with “Never” coded as zero intake. Blank or missing FFQ responses were treated as missing rather than zero intake. This distinction was central to the analysis because missingness can be outcome- or subgroup-dependent and may otherwise create artificial dietary structure^67,68^. For each participant, we calculated the number and proportion of missing FFQ items. Participants with more than 10% missing FFQ responses were excluded from the primary PCA analysis. Remaining missing values were median-imputed after the participant-level missingness filter. Food variables with insufficient nonmissing data or zero variance were removed before PCA. Total energy intake was winsorized at the 1st and 99th percentiles, log-transformed, and z-standardized.

### Dietary PCA in VDAART

Because FFQ-derived food-intake frequencies were sparse and right-skewed, we applied a log1p transformation to reduce the influence of extreme intake values while retaining zero intakes^69^. To identify food co-consumption structure independent of overall energy intake, each log-transformed food variable was residualized for log total energy. The residualized food matrix was standardized to mean 0 and SD 1, and PCA was performed on the standardized residuals, corresponding to analysis of the correlation matrix rather than the covariance matrix. Other demographic and clinical covariates were not removed before PCA; instead, they were adjusted for in downstream asthma/wheeze models. This approach preserves interpretable dietary variation while still estimating covariate-adjusted phenotype associations. The first 10 variance-ranked PCs were retained for visualization and downstream association analyses. Food items with absolute loadings ≥0.15 were considered major contributors for visualization and interpretation. Each dietary PC was interpreted primarily according to its major positive-loading foods, because higher PC scores indicate greater intake of these foods. Major negative-loading foods were also reported to describe the opposing dietary contrast captured by the component. PC signs were not oriented according to asthma/wheeze associations; instead, native PC scores were used in association models, and positive- and negative-loading foods were used only to interpret the dietary meaning of each PC. For multi-omic analyses, we used the selected PC scores as continuous dietary-pattern exposures and interpreted their microbiome and metabolomic associations in relation to the food-loading structure of each PC.

### Association between VDAART dietary PCs and asthma/wheeze

We tested associations between dietary PCs and year-3 asthma/wheeze using logistic regression. Individual PC models included a standardized PC score as the exposure and adjusted for child sex, study site, maternal education, child race, and gestational age. Odds ratios were reported per 1-standard-deviation higher dietary PC score. As the individual PC analyses were conducted as follow-up analyses to the global PC-set test and were used to identify which variance-ranked dietary axes contributed to the overall signal, we report nominal p values rather than using adjusted p values to make PC-level discovery claims. We also evaluated the joint contribution of PC1–PC10 using likelihood-ratio testing, comparing models with and without the PC block. Empirical significance was assessed by repeatedly permuting asthma/wheeze status and recalculating the likelihood-ratio statistic, thereby generating a null distribution for the association expected in the absence of a relationship between dietary PC structure and asthma/wheeze.

### NHANES external validation

We used the NHANES August 2021–August 2023 data as an external validation dataset^70^. NHANES is a nationally representative survey that combines interviews, physical examinations, and dietary assessment^70^. The primary NHANES validation sample included participants aged 2–3 years, with sensitivity analyses among participants aged 2–5 and 2–8 years. The primary NHANES outcome was current asthma, defined as ever having been told by a health professional that the participant had asthma and still having asthma at the time of interview, using MCQ010 and MCQ035.

NHANES dietary exposures were derived from the Day 1 Individual Foods file and Day 1 Total Nutrient Intakes file^71^. The Day 1 individual-food file provides one record per food consumed in the 24-hour recall period, while the total nutrient file provides total energy intake and dietary recall variables^71^. For PC1, PC3, and PC9, positive- and negative-loading food clusters were defined from the VDAART PCA loading structure. In VDAART, the native standardized PC score represents the positive-loading side of the axis, whereas the sign-reversed score represents the negative-loading side; these are mathematically complementary and yield identical statistical evidence with opposite effect directions. In NHANES, intake density was calculated separately for the positive- and negative-loading food clusters as total grams of foods in that cluster per 1000 kcal. Each cluster density was log1p-transformed and standardized, and the PC proxy score was defined as the standardized positive-cluster density minus the standardized negative-cluster density. The final contrast score was then standardized so that odds ratios corresponded to a 1-SD higher NHANES PC proxy score. To make effect estimates comparable across PCs, each proxy was then oriented to match the VDAART asthma/wheeze association direction. Thus, higher risk-aligned proxy scores represented the side of each VDAART dietary PC associated with higher asthma/wheeze risk. At the participant level, we calculated any intake, total grams, and grams per 1000 kcal separately for each food-cluster proxy. The primary NHANES exposure was log1p grams per 1000 kcal, z-standardized within each age-window-specific analytic sample. Secondary exposure definitions included any intake and log1p total grams. Survey-weighted logistic regression models were fitted for current asthma, adjusting for age, sex, race/ethnicity, poverty-income ratio, a missing-PIR indicator, and total energy intake. Models incorporated NHANES dietary weights, strata, and primary sampling units.

### Multi-omics data processing in VDAART

We analyzed three omics layers: gut microbiome, stool metabolome, and plasma metabolome. Stool and plasma samples were available for 340 children. Stool microbiome profiles were generated by 16S rRNA gene sequencing and included 209 microbial taxa after sequence processing and denoising using QIIME2/DADA2-based workflows^66^. Microbial taxa with a prevalence below 2% were excluded before association testing. Stool and plasma metabolomic profiles were measured using ultrahigh-performance liquid chromatography–mass spectrometry, with metabolite identification based on comparison to curated libraries of purified standards^66,72,73^. A total of 1,298 stool metabolites and 1,064 plasma metabolites were quantified. Microbiome features were transformed using sample-wise centered log-ratio transformation after adding a small pseudocount^74^. Stool and plasma metabolomics matrices were transformed using the same centered log-ratio framework for consistency with the compositional analysis pipeline^75^.

### Dietary pattern–omics association models

For each omics feature, we fitted a linear regression model:

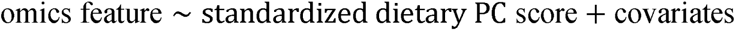

Models were adjusted for child sex, study site, maternal education, child race and gestational age. Omics features with insufficient complete-case sample size or zero variance were excluded. For each omics layer and dietary exposure, P values were adjusted using the Benjamini–Hochberg false discovery rate procedure^76^. Results were summarized by the number of nominal associations, FDR <0.10 associations, and FDR <0.05 associations. For visualization, the top-ranked features in each omics layer were plotted using signed −log10(P), where positive values indicate higher feature abundance with higher dietary PC score and negative values indicate lower feature abundance with higher PC1-negative food-cluster score.

### Statistical analysis

All analyses were performed in R. Logistic regression was used for binary asthma/wheeze outcomes in VDAART. NHANES validation models used survey-weighted logistic regression to account for complex sampling design. Linear regression was used for feature-wise omics association testing. Continuous exposures were z-standardized, and odds ratios were reported per 1-standard-deviation increase. Statistical significance was assessed using two-sided tests. For omics analyses, false discovery rate was controlled using the Benjamini– Hochberg procedure.

## Results

### Data-driven dietary patterns capture food co-consumption structure in early childhood

We first used energy-adjusted food-level PCA to summarize age-3 dietary intake patterns in VDAART. This data-driven approach identified latent co-consumption axes that reflect how foods were consumed together, rather than evaluating individual foods in isolation. For each PC, we visualized food items with an absolute loading ≥0.15 to focus on foods making the strongest contributions to each dietary axis (see **Figure 2**). The first 10 PCs accounted for 36.85% of the total variance and captured distinct food co-consumption structures, including patterns involving fruits and vegetables, sweetened snacks, fried foods, processed meats, dairy, breads/cereals, fish, and other commonly consumed foods. Notably, PC1 represented a major confectionery food such as cakes and pies. The negative-loading side of PC1 included tomatoes, carrots, spinach, cantaloupe, mixed vegetables, pear, berries, and apple/applesauce, whereas the positive-loading side included chips, fruit drinks, cake/cupcake, donuts/fried dough, sweet rolls/muffins, pie, and french fries. PC2 captured a separate mixed meat/vegetable dietary axis, with higher loadings for foods such as ham/baked steak, pork, cabbage/coleslaw/cauliflower, sweet potatoes/yams, and canned tuna. PC3 was driven by meat products captured a different contrast with positive-loading side characterized by processed meats and fried foods, including hot dogs, fried chicken/chicken nuggets, sausage, bacon, and cold cuts, whereas the negative-loading side included chocolate candy, cookies/brownies, cornbread/tortilla, pie, cream cheese, cheese, cake/cupcake, other candy, butter, dark bread, and yogurt. The loading structure highlighted the major food combinations underlying each dietary PC and provided the interpretive framework for relating PC scores to asthma/wheeze, NHANES food-cluster proxies, and multi-omic signatures.

**Figure 2.**
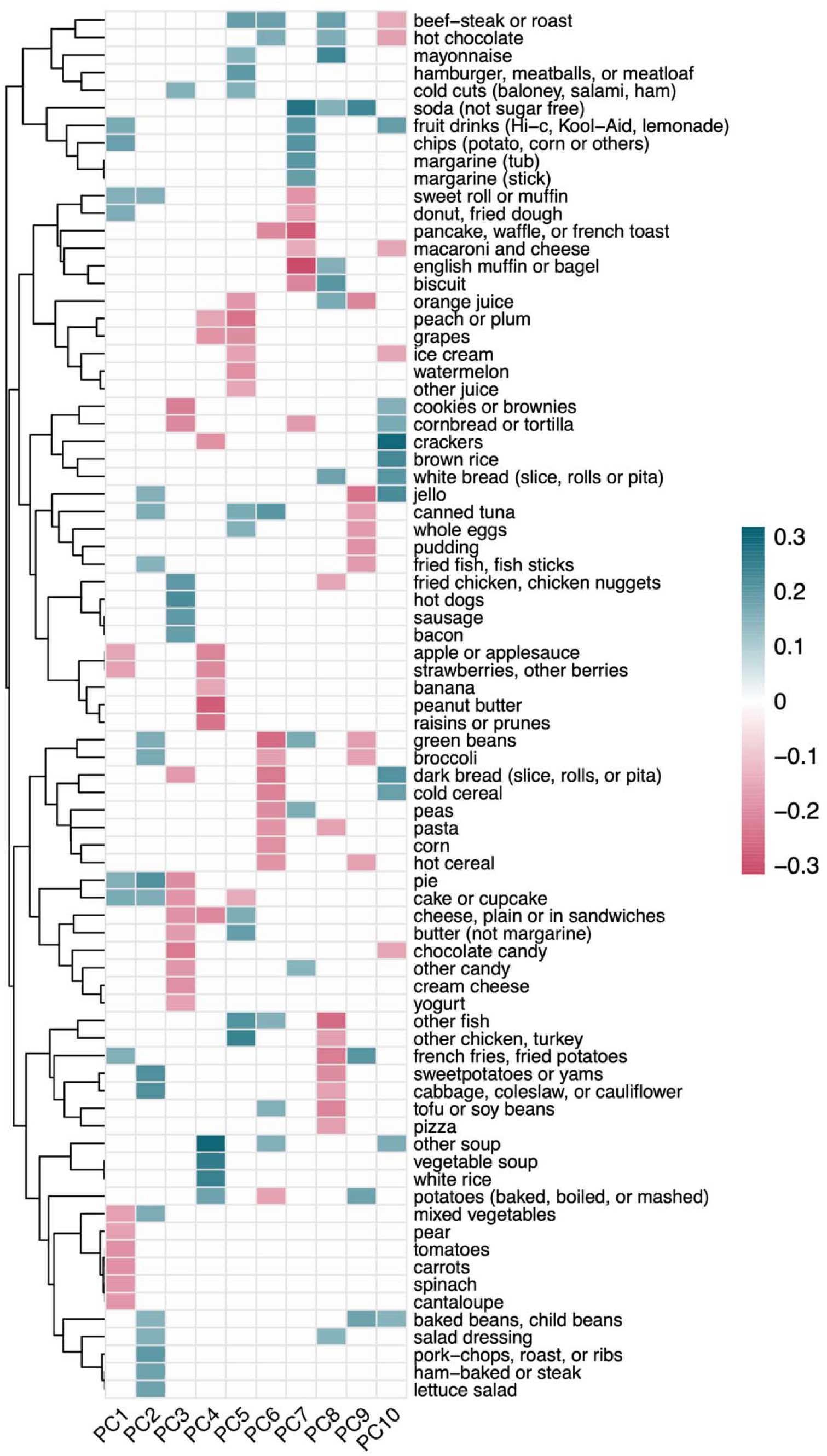
Energy-adjusted dietary principal components identify distinct food co-consumption patterns. Heatmap of food loadings for the first 10 dietary principal components derived from 87 age-3 VDAART food frequency questionnaire items after energy adjustment. Rows represent food items and columns represent PCs. To improve interpretability, only food– PC loadings with an absolute loading magnitude ≥ 0.15 are displayed. Color indicates the direction and magnitude of the loading.

### Dietary PCs are jointly associated with asthma/wheeze in VDAART

We then tested whether the dietary-pattern scores, considered together, improved the prediction of asthma/wheeze beyond the covariates alone. To do this, we compared two logistic regression models: a base model containing the adjustment covariates only, and an expanded model containing the same covariates plus the first 10 dietary PC scores. The improvement in model fit was summarized by a likelihood-ratio statistic. To determine whether this improvement was larger than expected by chance, we repeated the same analysis after randomly permuting the asthma/wheeze outcome and generated an empirical null distribution. The observed likelihood-ratio statistic was 34.2, which was greater than almost all permuted values, corresponding to an empirical p value of 2 10 (see **Figure 3a**). Thus, the dietary PC scores jointly contributed asthma/wheeze-related information beyond the adjustment covariates.

**Figure 3.**
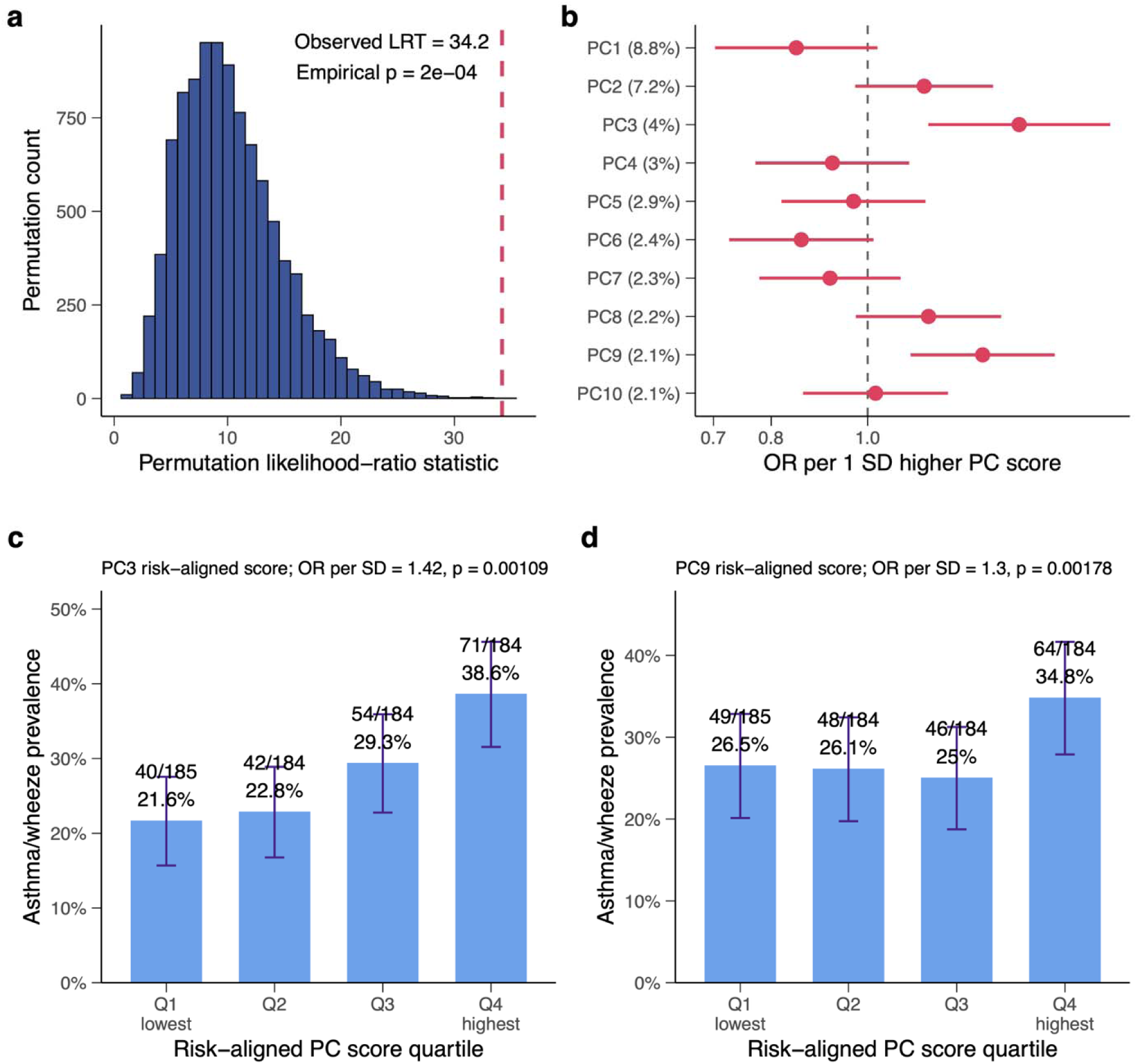
Dietary principal components are jointly associated with asthma/wheeze in VDAART. **(a)** Permutation-based global test of the dietary PC set. The observed likelihood-ratio statistic was 34.2, exceeding the empirical null distribution, with empirical p = 2 × 10□□. **(b)** Odds ratios and 95% confidence intervals for asthma/wheeze per 1-SD higher PC score across PC1–PC10. PC3 showed the strongest positive association with asthma/wheeze. **(c)** Asthma/wheeze prevalence across quartiles of the PC3 risk-aligned score. **(d)** Asthma/wheeze prevalence across quartiles of the PC9 risk-aligned score. Risk-aligned scores were oriented so that higher values correspond to higher estimated asthma/wheeze risk.

After establishing that the dietary PC set was associated with asthma/wheeze, we next identified which individual dietary axes contributed most strongly to this overall signal. Each PC was tested separately in covariate-adjusted logistic regression models to estimate the association between a 1-SD higher PC score and asthma/wheeze. Among individual PCs, PC3 showed the strongest positive association with asthma/wheeze (see **Figure 3b**). Each 1-SD higher PC3 score was associated with increased odds of asthma/wheeze, and the risk-aligned PC3 score showed a graded prevalence pattern across quartiles. Asthma/wheeze prevalence increased from 21.6% in the lowest quartile to 38.6% in the highest quartile (see **Figure 3c**). PC9 also showed a positive association after risk alignment, with asthma/wheeze prevalence increasing from 26.5% in Q1 to 34.8% in Q4 (see **Figure 3d**). Although PC1 was not the strongest asthma-associated PC in VDAART, it represented a biologically interpretable and externally testable dietary axis. These results suggest that childhood asthma/wheeze is associated with multivariate dietary-pattern structure rather than a single isolated food item.

### Age-3 dietary patterns were prospectively associated with asthma/wheeze at age 6

To evaluate whether age-3 dietary patterns were associated with a later and more clinically stable respiratory phenotype, we repeated the dietary PC analysis using asthma/wheeze diagnosis at age 6 as the outcome. This analysis included 791 children with available age-6 outcome data and diet data, including 394 with asthma/wheeze and 397 controls. The age-3 dietary PC set showed evidence of prospective association with age-6 asthma/wheeze. In the global likelihood-ratio test, adding the dietary PC block improved model fit compared with the covariate-only model, with an observed likelihood-ratio statistic of 20.3 and an empirical permutation p value of 0.033 (see **Fig.S1a**). Individual PC analyses showed broadly consistent directions with the age-3 asthma/wheeze results, supporting the stability of the dietary-pattern signal across outcome definitions. The strongest positive associations with age-6 asthma/wheeze were observed for PC2 and PC9 (see **Fig.S1b**). For PC2, each 1-SD higher risk-aligned score was associated with higher odds of age-6 asthma/wheeze, and asthma/wheeze prevalence increased across quartiles from 42.2% in Q1 to 52.2% in Q4 (see **Fig.S1c**). PC9 showed a similar positive association, with prevalence highest in the top risk-aligned quartile: 52.7% in Q4 compared with 46.5%, 43.5%, and 41.8% in Q1–Q3, respectively (see **Fig.S1d**). These prospective analyses indicate that dietary patterns measured at age 3 were associated with asthma/wheeze status at age 6, strengthening the evidence that the observed diet–asthma relationships were not limited to cross-sectional associations at the time of dietary assessment. The consistency in overall PC association directions between the age-3 and age-6 outcomes further supports the robustness of the dietary-pattern signal.

### NHANES provides directional external support for the PC dietary axis

We next evaluated whether the VDAART PC dietary axis could be approximated in an age-matched external population using NHANES Day 1 dietary recall data. The PC3-positive proxy captured the meat product side of PC3, e.g., hot dogs, sausage, and bacon, while the PC3-negative proxy captured the dairy product side (see **SI Table 1**). The proxy score was defined as positive-loading cluster intake density minus negative-loading cluster intake density. For each food-cluster proxy, we constructed three complementary exposure measures: any intake, a binary indicator of whether any cluster food was reported; amount, the log-transformed total grams of cluster foods consumed; and density, the log-transformed grams of cluster foods per 1000 kcal. The primary analysis focused on children aged 2–3 years to match the VDAART age-3 dietary assessment and included 168 children, of whom 10 had current asthma (see **Figure 4a & b**). We then examined nested age-window sensitivity analyses among children aged 2–5 years and 2–8 years. Accordingly, the age-window analysis evaluates whether the density-based association was stable across broader pediatric samples, whereas the exposure-definition analysis evaluates whether results were sensitive to using binary intake, absolute amount, or energy-adjusted density.

**Figure 4.**
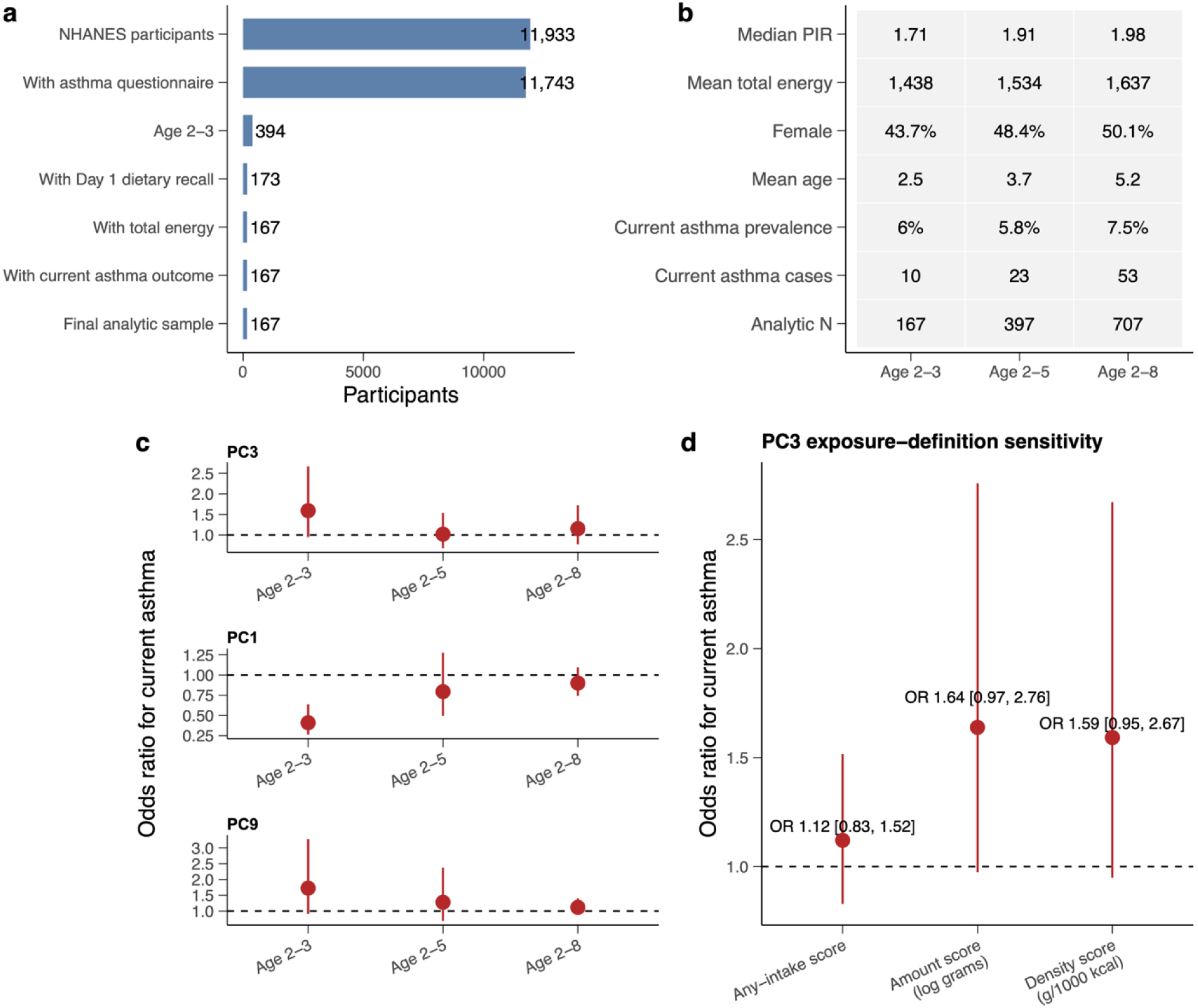
External NHANES validation of the VDAART PC food-cluster proxies. **(a)** NHANES analytic sample flow for the pediatric validation analysis. Participants were restricted to children with asthma questionnaire data, age eligibility, Day 1 dietary recall, total energy data, and current asthma outcome information. **(b)** Characteristics of analytic samples across the age 2–3, 2–5, and 2–8 window. **(c)** Association between PC food-cluster proxy density scores and current asthma across age windows. Each proxy score was defined as positive-loading cluster intake density minus negative-loading cluster intake density; PC3 was the main proxy, with PC1 and PC9 shown as supplementary analyses. PC proxy scores were risk-aligned according to th VDAART asthma/wheeze association direction, so odds ratios greater than 1 indicate higher current asthma odds for the VDAART risk-associated side of each dietary axis. **(d)** Sensitivity analysis for the main PC3 proxy using alternative exposure definitions: any intake, log-transformed grams, and log-transformed grams per 1000 kcal.

**Figure 5.**
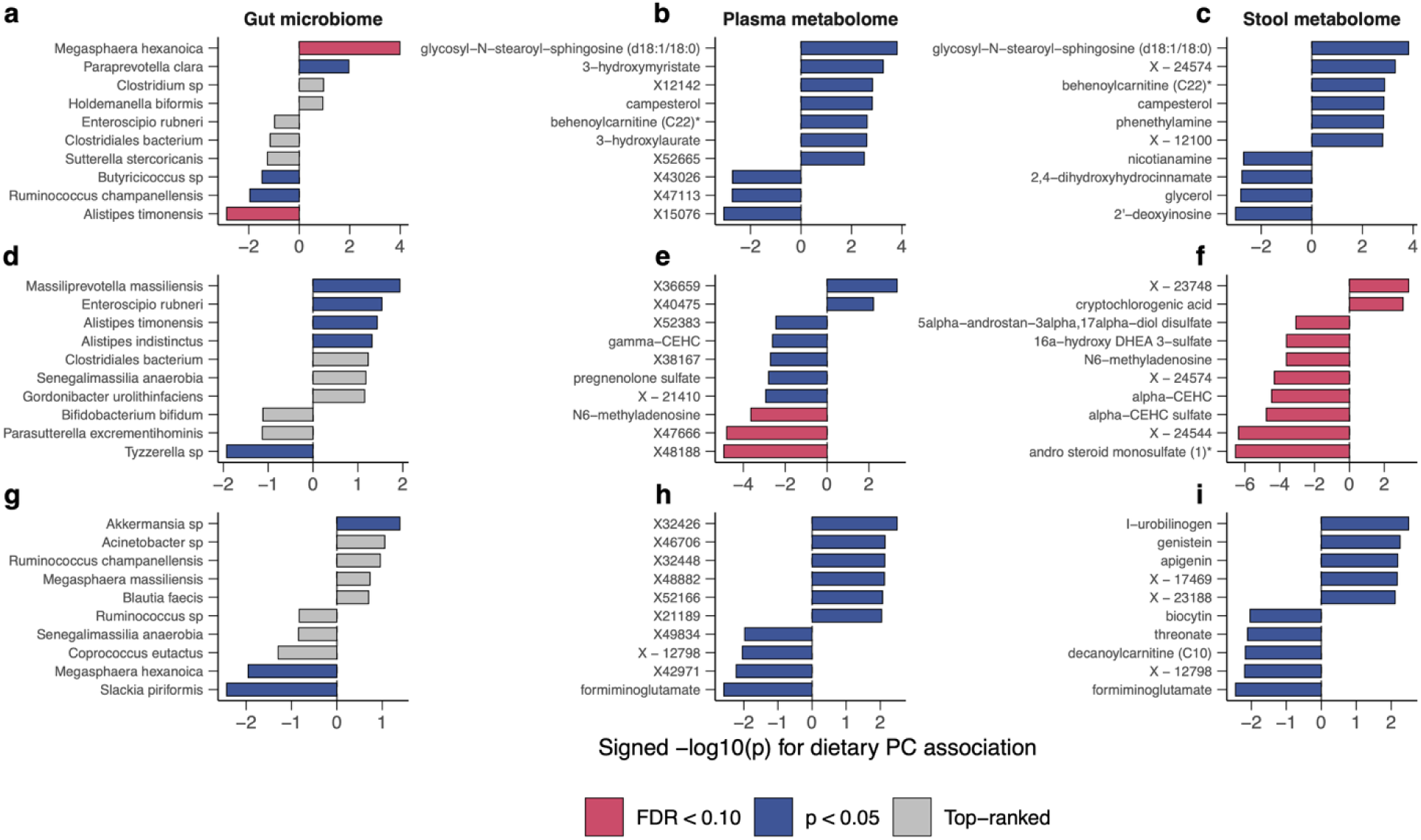
PC1 and PC3 and PC9 dietary patterns are associated with microbial and metabolomic features. Top-ranked gut microbiome, plasma metabolome, and stool metabolome features associated with standardized PC1, PC3 and PC9 dietary-pattern scores. Panels a–c show Associations between the PC1 (a-c), PC3 (d-f), PC9 (g-i) dietary-pattern score and gut microbiome, plasma metabolome, and stool metabolome features. Bars show signed −log10(p), with positive values indicating higher abundance with the dietary PC and negative value indicating lower abundance. Red bars indicate FDR < 0.10, blue bars indicate nominal p < 0.05, and gray bars indicate top-ranked features not meeting nominal significance.

In the primary age 2–3 analysis, higher intake density of the PC3 food-cluster proxy was associated with higher odds of current asthma (OR per 1-SD increase, 1.59; 95% CI, 0.95–2.67; see **Figure 4c**). The total-amount definition showed a similar association (OR, 1.64; 95% CI, 0.97–2.76), whereas the any-intake estimate was less strong (OR, 1.12; 95% CI, 0.83–1.52; **Figure 4d**). The OR for intake density was 1.02 (95% CI, 0.64–1.54) at ages 2–5 years, and 1.15 (95% CI, 0.77–1.72) at ages 2–8 years (see **Figure 4d**). Exploratory NHANES proxies for PC1 and PC9 were also evaluated. The OR for intake density was 0.41 (95% CI, 0.26–0.64) for PC1, and 1.73 (95% CI, 0.91–3.28) for PC9 (see **Figure 4c**). These findings provide age-matched external support in NHANES for the direction of the VDAART dietary-pattern signal, although they should be interpreted as supportive validation rather than strict replication because NHANES used 24-hour dietary recall rather than the original VDAART FFQ-derived PC score.

### PC1, PC3, and PC9 dietary patterns are associated with distinct microbial and metabolomic profiles

We finally examined whether the prioritized dietary axes were associated with gut microbiome, stool metabolome, and plasma metabolome features. We found that PC1 was associated with a cross-compartment lipid metabolic signature in both stool and plasma. For example, PC1-associated metabolites included glycosyl-N-stearoyl-sphingosine, behenoylcarnitine C22, 3-hydroxylaurate, 3-hydroxymyristate, campesterol, glycerol, and related lipid or sterol metabolites. Sphingolipid metabolism is relevant to asthma biology because disrupted ceramide regulation can enhance allergic airway inflammation, whereas sphingosine-1-phosphate can promote airway smooth-muscle hyperresponsiveness and proliferation^77,78^. Several neonatal acylcarnitines have also been associated with subsequent early-childhood wheeze^79^, although the asthma-specific role of behenoylcarnitine C22 remains unknown. This pattern suggests that the PC dietary axis is linked to sphingolipid-related metabolism, fatty-acid transport and energy metabolism, hydroxy fatty acid metabolism, sterol metabolism, and another lipid-backbone metabolism. These associations place PC1 within lipid-metabolic domains relevant to airway inflammation and airway smooth-muscle function, without establishing a causal pathway or clinically validated biomarker.

PC3 showed a partially distinct metabolomic profile. PC3-associated metabolites included alpha-CEHC, alpha-CEHC sulfate, gamma-CEHC, pregnenolone sulfate, DHEA-related steroid sulfates, andro-steroid sulfates, N6-methyladenosine, 2 ′ -deoxyinosine, N6-formyllysine, trans-4-hydroxyproline, and cryptochlorogenic acid. Alpha-CEHC and gamma-CEHC are downstream metabolites of alpha- and gamma-tocopherol, respectively^80^. Because tocopherol isoforms can exert divergent immunoregulatory effects during allergic inflammation^81,82^, these signals are best interpreted as reflecting vitamin E metabolism rather than evidence that vitamin E is uniformly protective. DHEA-S-related steroid metabolism has been linked to asthma control and lung function in some adult studies^83,84^. Nucleoside-related signals may also be relevant because adenosine and extracellular ATP signaling can contribute to bronchoconstriction, inflammatory signaling, and type 2 inflammatory responses in the airway epithelium^85–87^. Hydroxyproline is related to collagen metabolism and has been linked to airway remodeling in experimental studies^88^, although stool or circulating trans-4-hydroxyproline is not a direct measure of airway fibrosis. These metabolites suggest biological domains involving vitamin E metabolism, oxidative stress-related processes, steroid hormone metabolism and immune regulation, nucleoside-related metabolism, and tissue remodeling.

PC9 showed a third, less pronounced microbial and metabolomic profile. Among the named stool metabolites, higher PC9 scores were associated with higher levels of l-urobilinogen, genistein, and apigenin and with lower levels of biocytin, threonate, decanoylcarnitine C10, and formiminoglutamate. For PC9, the strongest microbiome and metabolomic associations in the full feature-wise analysis were nominal, with no features reaching FDR <0.10 after multiple-testing correction. Together, the parallel microbial and metabolomic results suggest that the asthma-associated dietary PCs are not merely statistical food-pattern summaries. Rather, they are associated with biologically interpretable metabolic domains that are plausibly relevant to asthma, including airway inflammation, oxidative stress, lipid signaling, and tissue remodeling.

## Discussion

In this study, we applied food-level PCA to identify early-childhood dietary co-consumption patterns and tested their associations with asthma/wheeze. The main finding is that asthma/wheeze was associated with multivariate dietary-pattern structure rather than a single isolated food. The global PC association test was significant, and PC3 showed a clear graded relationship with asthma/wheeze prevalence. Food-cluster proxies representing both sides of PC1 were externally evaluated in NHANES and showed directionally consistent associations with current asthma. Finally, parallel microbial and metabolomic analyses linked PC1, PC3 and PC9 to distinct and biologically interpretable profiles.

Dietary exposures are highly correlated, e.g., children who consume one food often consume related foods as part of a broader eating pattern. Therefore, single-food analyses can be unstable or misleading if they do not account for co-consumption structure. The PCA approach allowed us to identify dietary axes that reflect real-world food combinations^89^. This is especially important for pediatric dietary data, where reporting variation, portion-size uncertainty, and sparse intake for some foods can complicate individual-food inference^43^.

The global permutation result supports the idea that the dietary-pattern space as a whole carries asthma/wheeze-related information. PC3 emerged as the strongest VDAART asthma-associated axis, whereas PC1 provided a major interpretable food-pattern axis that could be tested in NHANES. This distinction is important: PC3 appears most relevant for VDAART asthma association, while PC1 provides the strongest external validation opportunity because its food clusters representing both sides of the axis could be translated into NHANES dietary recall variables. The omics results suggest that PC1 and PC3 may represent different diet-linked biological routes. PC1 was most strongly linked to lipid-related metabolites across stool and plasma, including sphingolipid-related metabolites, long-chain acylcarnitines, hydroxy fatty acids, and sterol-related metabolites. Sphingolipid biology has been repeatedly connected to asthma through airway inflammation^77^, airway smooth muscle dysfunction^78^, epithelial biology^90^, and airway remodeling^91^; altered sphingolipid metabolism has also been associated with asthma in metabolomic studies^92,93^. PC3 showed a more diverse metabolic profile involving vitamin E/tocopherol metabolites, steroid sulfates, nucleoside-related metabolites, and hydroxyproline-related features. The CEHC metabolites provide a useful interpretation bridge: alpha-CEHC and gamma-CEHC are downstream markers of tocopherol/vitamin E metabolism, and vitamin E isoforms have been studied in allergic inflammation and asthma biology^94^. PC9 yielded mainly nominal associations, including stool signals related to plant-derived flavonoids and intermediary metabolism; these findings remain exploratory and require independent confirmation.

The steroid sulfate findings are also biologically plausible. DHEA/DHEA-S and related steroid pathways have been connected to asthma outcomes^83^ and lung function^84^, including work suggesting that DHEA biology may be relevant to asthma outcomes in specific subgroups^95^. Nucleoside-related metabolites should be interpreted cautiously, but they are relevant to airway biology because adenosine and purinergic signaling can influence bronchoconstriction and airway obstruction^96^, inflammatory signaling^85,86^, and epithelial stress responses^87^. Hydroxyproline-related signals may reflect collagen turnover or tissue remodeling^88^, a process central to chronic airway disease^97^; emerging metabolomics literature also supports amino-acid and remodeling-related metabolic changes in asthma models and pediatric asthma contexts^98–100^. Several of the metabolomic signals observed here overlap with pathways previously implicated in VDAART and pediatric asthma metabolomics studies, supporting the biological relevance of the dietary-pattern associations. For example, lipid-related metabolites^57,93^, oxidative-stress-associated pathways^101^, and amino-acid-related metabolic features^98^ have been reported in prior VDAART or pediatric asthma analyses. In contrast, other signals highlighted in the present analysis appear to extend beyond previously reported VDAART findings. These include dietary PC-associated steroid sulfate metabolites, nucleoside-related metabolites, sphingolipid-related metabolites, long-chain acylcarnitines, and hydroxyproline-related signals, which may reflect endocrine-immune regulation, purinergic or epithelial stress biology, lipid remodeling, fatty-acid oxidation, and tissue-remodeling processes. Thus, the final multi-omic analysis both recapitulates known asthma-relevant metabolic domains and identifies additional stool and plasma metabolomic features linked specifically to asthma-associated dietary co-consumption patterns.

A major strength of this analysis is the integration of data-driven dietary patterns, external population-level validation, and multi-omic biological annotation. The use of PCA reduces overreliance on individual foods and better reflects co-consumption structure. The NHANES analysis provides an independent check on food-cluster proxies representing both sides of an interpretable PC1 axis, and the metabolomics results help translate dietary PCs into biological pathways. Several limitations should be acknowledged. First, the analysis is observational and cannot establish causality. Second, dietary data are subject to measurement error, especially in young children^44^. Third, the NHANES validation used dietary recall data and a current asthma outcome, which are not identical to the VDAART exposure and outcome structure. Fourth, some metabolite features are unannotated, and unknown IDs should not be biologically interpreted^102,103^. Fifth, microbiome taxon-level signals are exploratory and can be affected by sparsity and count-distribution properties^104^, compositionality^105^, and technical variation across laboratory and bioinformatic workflows^106^. Finally, the metabolite pathway interpretation is hypothesis-generating and requires validation using targeted metabolomics, independent cohorts, and ideally longitudinal or interventional designs.

## Data Availability

Data described in the manuscript, code book, and analytic code will be made available upon request, pending application and approval.

## Acknowledgements

X.W.W. acknowledges the funding support from National Institutes of Health (K25HL166208).

## Author contributions

X.W.W. conceived and designed the project. Y.W. performed data analysis. Y.W. and X.W.W. wrote the manuscript. Y.W. and X.W.W., T.C., S.T.W., and Y.Y.L. interpreted the results. All authors approved the manuscript.

---

**Figure S1.**
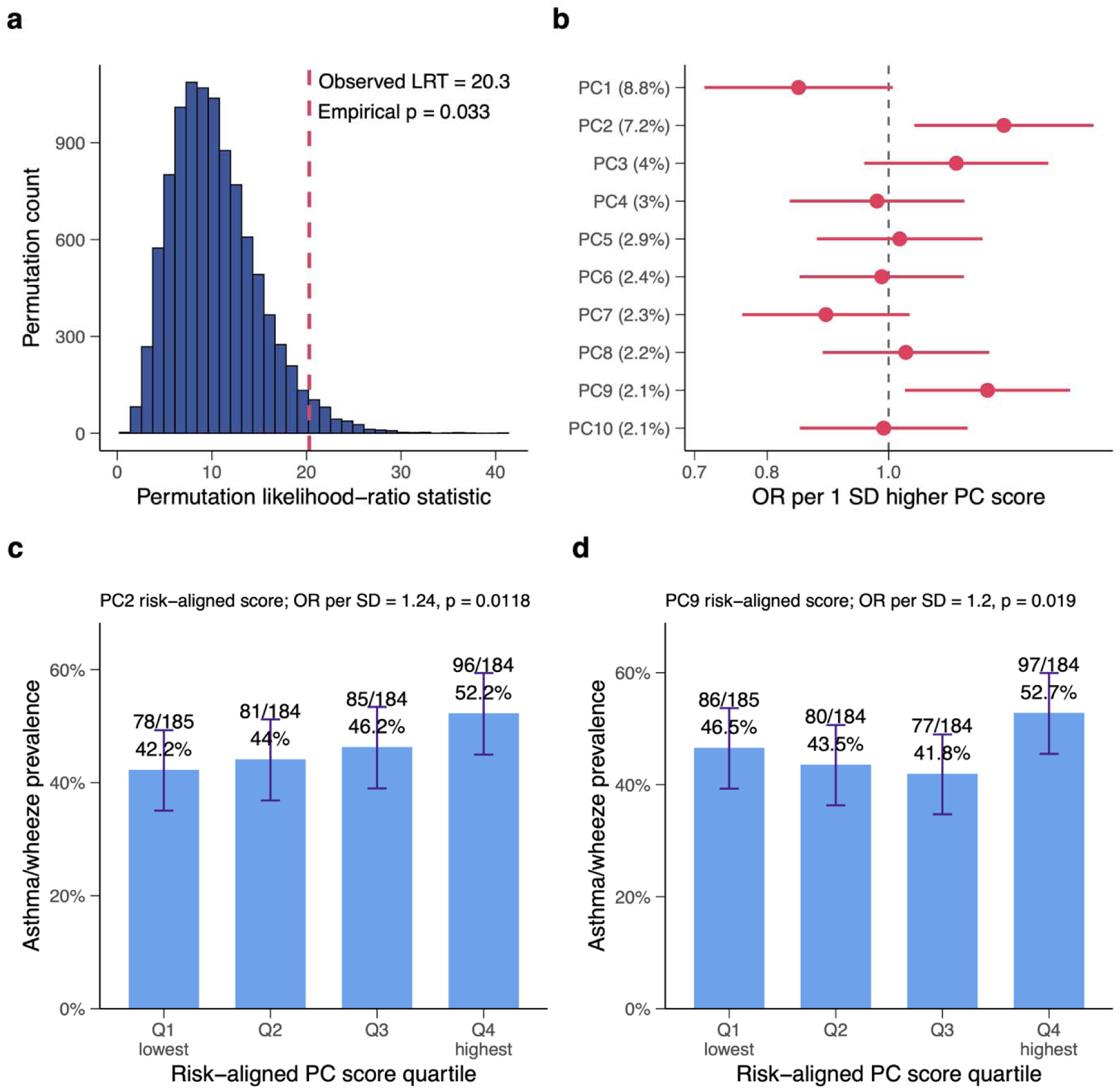
Dietary principal components are jointly associated with asthma/wheeze in VDAART at year 6. **(a)** Permutation-based global test of the dietary PC set. The observed likelihood-ratio statistic was 20.3, exceeding the empirical null distribution, with empirical p = 0.033. **(b)** Odds ratios and 95% confidence intervals for asthma/wheeze per 1-SD higher PC score across PC1–PC10. PC2 showed the strongest positive association with asthma/wheeze. **(c)** Asthma/wheeze prevalence across quartiles of the PC2 risk-aligned score. **(d)** Asthma/wheeze prevalence across quartiles of the PC9 risk-aligned score. Risk-aligned scores were oriented so that higher values correspond to higher estimated asthma/wheeze risk.

**Supplementary Table 1.**
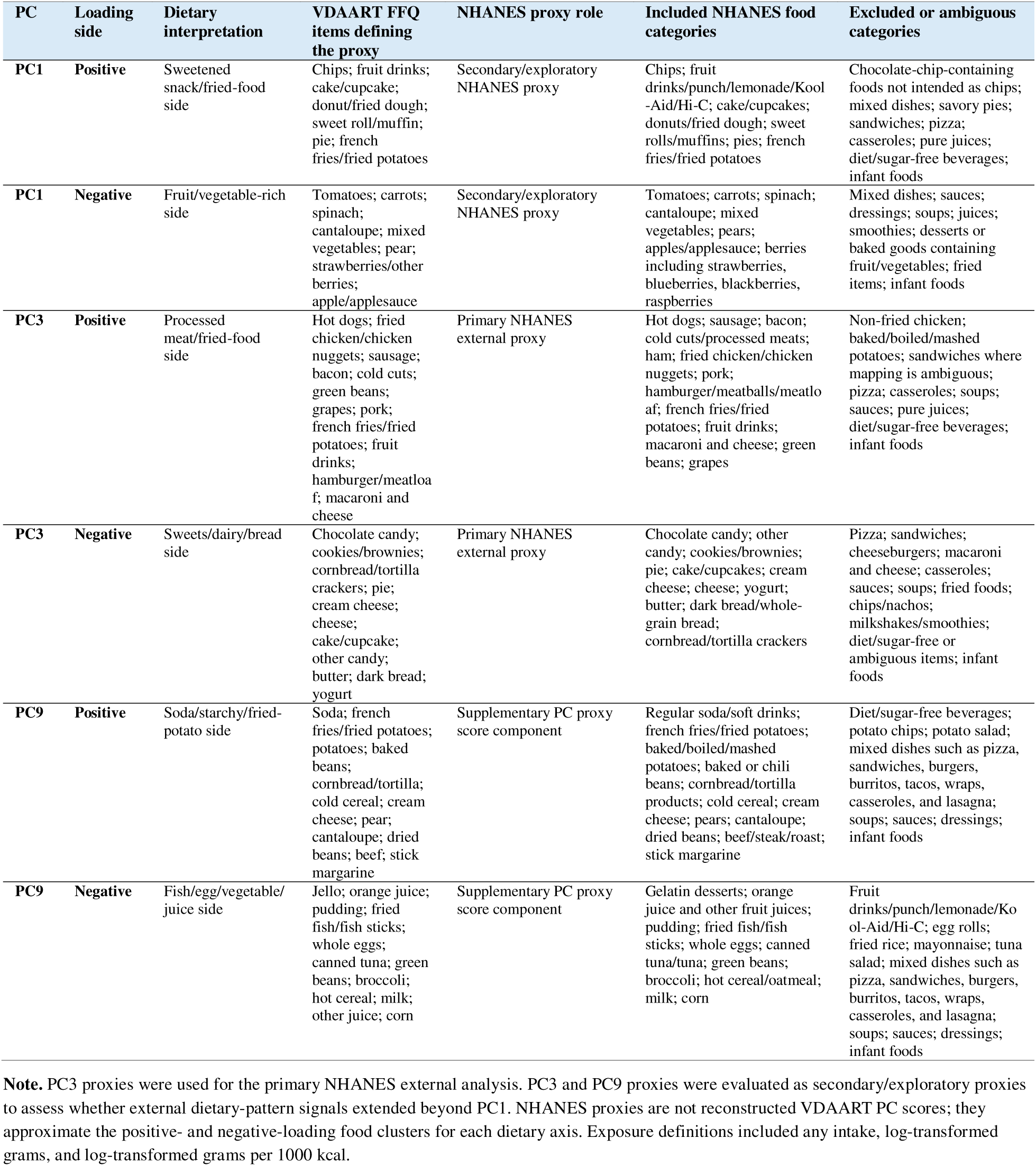
VDAART PCA-derived food clusters used to construct NHANES dietary proxies. Food clusters were defined from VDAART FFQ items with prominent PCA loadings and then mapped conservatively to NHANES Day 1 dietary recall food codes.

| PC | Loading side | Dietary interpretation | VDAART FFQ items defining the proxy | NHANES proxy role | Included NHANES food categories | Excluded or ambiguous categories |
| --- | --- | --- | --- | --- | --- | --- |
| PC1 | Positive | Sweetened snack/fried-food side | Chips; fruit drinks; cake/cupcake; donut/fried dough; sweet roll/muffin; pie; french fries/fried potatoes | Secondary/exploratory NHANES proxy | Chips; fruit drinks/punch/lemonade/Kool-Aid/Hi-C; cake/cupcakes; donuts/fried dough; sweet rolls/muffins; pies; french fries/fried potatoes | Chocolate-chip-containing foods not intended as chips; mixed dishes; savory pies; sandwiches; pizza; casseroles; pure juices; diet/sugar-free beverages; infant foods |
| PC1 | Negative | Fruit/vegetable-rich side | Tomatoes; carrots; spinach; cantaloupe; mixed vegetables; pear; strawberries/other berries; apple/applesauce | Secondary/exploratory NHANES proxy | Tomatoes; carrots; spinach; cantaloupe; mixed vegetables; pears; apples/applesauce; berries including strawberries, blueberries, blackberries, raspberries | Mixed dishes; sauces; dressings; soups; juices; smoothies; desserts or baked goods containing fruit/vegetables; fried items; infant foods |
| PC3 | Positive | Processed meat/fried-food side | Hot dogs; fried chicken/chicken nuggets; sausage; bacon; cold cuts; green beans; grapes; pork; french fries/fried potatoes; fruit drinks; hamburger/meatloaf; macaroni and cheese | Primary NHANES external proxy | Hot dogs; sausage; bacon; cold cuts/processed meats; ham; fried chicken/chicken nuggets; pork; hamburger/meatballs/meatloaf; french fries/fried potatoes; fruit drinks; macaroni and cheese; green beans; grapes | Non-fried chicken; baked/boiled/mashed potatoes; sandwiches where mapping is ambiguous; pizza; casseroles; soups; sauces; pure juices; diet/sugar-free beverages; infant foods |
| PC3 | Negative | Sweets/dairy/bread side | Chocolate candy; cookies/brownies; cornbread/tortilla crackers; pie; cream cheese; cheese; cake/cupcake; other candy; butter; dark bread; yogurt | Primary NHANES external proxy | Chocolate candy; other candy; cookies/brownies; pie; cake/cupcakes; cream cheese; cheese; yogurt; butter; dark bread/whole-grain bread; cornbread/tortilla crackers | Pizza; sandwiches; cheeseburgers; macaroni and cheese; casseroles; sauces; soups; fried foods; chips/nachos; milkshakes/smoothies; diet/sugar-free or ambiguous items; infant foods |
| PC9 | Positive | Soda/starchy/fried-potato side | Soda; french fries/fried potatoes; potatoes; baked beans; cornbread/tortilla; cold cereal; cream cheese; pear; cantaloupe; dried beans; beef; stick margarine | Supplementary PC proxy score component | Regular soda/soft drinks; french fries/fried potatoes; baked/boiled/mashed potatoes; baked or chili beans; cornbread/tortilla products; cold cereal; cream cheese; pears; cantaloupe; dried beans; beef/steak/roast; stick margarine | Diet/sugar-free beverages; potato chips; potato salad; mixed dishes such as pizza, sandwiches, burgers, burritos, tacos, wraps, casseroles, and lasagna; soups; sauces; dressings; infant foods |
| PC9 | Negative | Fish/egg/vegetable/juice side | Jello; orange juice; pudding; fried fish/fish sticks; whole eggs; canned tuna; green beans; broccoli; hot cereal; milk; other juice; corn | Supplementary PC proxy score component | Gelatin desserts; orange juice and other fruit juices; pudding; fried fish/fish sticks; whole eggs; canned tuna/tuna; green beans; broccoli; hot cereal/oatmeal; milk; corn | Fruit drinks/punch/lemonade/Kool-Aid/Hi-C; egg rolls; fried rice; mayonnaise; tuna salad; mixed dishes such as pizza, sandwiches, burgers, burritos, tacos, wraps, casseroles, and lasagna; soups; sauces; dressings; infant foods |

**Note.** PC3 proxies were used for the primary NHANES external analysis. PC3 and PC9 proxies were evaluated as secondary/exploratory proxies to assess whether external dietary-pattern signals extended beyond PC1. NHANES proxies are not reconstructed VDAART PC scores; they approximate the positive- and negative-loading food clusters for each dietary axis. Exposure definitions included any intake, log-transformed grams, and log-transformed grams per 1000 kcal.

## Notes

### Competing Interest Statement

The authors have declared no competing interest.

### Clinical Trial

NCT00920621 (VDAART)

### Author Declarations

VDAART IRB approval was obtained from each of the three clinical centers and the Data Coordinating Center, that is, Washington University in St. Louis, Kaiser Health Care San Diego, Boston Medical Center and Brigham and Women's Hospital in Boston. Study subjects provided written, informed consent.

